# Evaluation of Large Language Models for Post-Cystectomy Sexual Health Counseling in Women

**DOI:** 10.64898/2026.06.25.26356154

**Authors:** Faheed Shafau, Aakash Dave, Ibukunoluwa Omole, Taeris Guzman, Najibah Rehman, Ekene Enemchukwu, Larissa Bresler

**Author notes:** **Corresponding Author:** Faheed Shafau, Michigan State University, College of Human Medicine, Grand Rapids, MI 49503, United States. These authors contributed equally to this work (Faheed Shafau, Aakash Dave). **Ethics and Consent:** This study was reviewed by the Michigan State University Human Research Protection Program and was determined not to involve human subjects as defined by federal regulations. Therefore, Institutional Review Board approval and informed consent were not required. **Conflicts of Interest:** The authors declare no conflicts of interest. **Funding Statement:** This research received no external funding. **Data availability statement:** All data analyzed are included in this manuscript and its supplementary information. **Artificial Intelligence Generated Content Declaration:** Artificial intelligence was used to generate patient educational materials. ChatGPT-4.2, Gemini 1.5, and Perplexity were used for the generation of data in the context of assessing quality of information. Artificial Intelligence was not used for the writing of this manuscript. All AI-assisted outputs were reviewed, verified, and edited by the authors, who take full responsibility for the content of the manuscript.

## Abstract

**Objective:** To evaluate the adherence to guidelines and readability of large language model-generated sexual health information related to female sexual dysfunction following cystectomy, and to determine whether adherence differs across models and prompt formats. A secondary objective was to introduce an analytic strategy using principal component analysis to examine the dimensions of readability metrics.

**Methods:** Three large language models (LLMs), ChatGPT, Gemini, and Perplexity were prompted with six clinical questions related to sexual function after cystectomy. Questions were phrased in long-form and short-form language. Responses were independently graded by two reviewers, derived from guideline recommendations. Linear mixed-effects models predicted adherence as functions of LLM, prompt, and reviewer, with clinical questions as a random intercept. Readability was assessed using five metrics, and principal component analysis (PCA) was used to determine latent structure.

**Results:** ChatGPT demonstrated the highest (estimated marginal mean [emm] = 0.769), outperforming Gemini (0.499) and Perplexity (0.457). Shorter, less complex prompts elicited higher adherence than more complex, clinical prompts. All models produced content that exceeded recommended reading levels. PCA demonstrated that a single dominant component accounted for 76.7% of variance across readability indices, indicating a shared underlying construct.

**Conclusion:** ChatGPT produced the most guideline-concordant information overall. High linguistic complexity was seen across models, highlighting a barrier to patient comprehension. These findings characterize large language models as variable medical information systems whose outputs rely heavily on prompt structure and model type.

## I. Introduction

Muscle-invasive bladder cancer (MIBC) is less common in women than in men, but women are more likely to present with late-stage disease and have poorer survival rates.^1^ A systematic review and meta-analysis of 69,666 patients found that women had a 23% higher risk of cancer-specific mortality (HR 1.23, 95% CI 1.15–1.31) and an 8% higher risk of overall mortality (HR 1.08, 95% CI 1.03–1.12) following radical cystectomy.^2^ Additionally, sexual dysfunction is highly prevalent among women with muscle-invasive bladder cancer, particularly following radical cystectomy.^3^ Sexual dysfunction following cystectomy extends beyond physiologic impairment, contributing to psychological distress, altered self-image, and long-term burden.

Sexual dysfunction following radical cystectomy has substantial psychosocial consequences beyond anatomic and functional changes. Women report distress related to body image, emotional well-being, and intimate relationships.^4^ Despite the prevalence and clinical significance of these concerns, sexual health counseling remains inconsistently delivered, with female patients less likely than their male counterparts to receive provider-initiated discussions addressing sexual function, psychological well-being, and survivorship needs.^5,6^ This literature shows that postoperative sexual health in women is closely linked to mental health, psychosocial support, and access to counseling domains that intersect with AI-mediated patient education tools. From a medical systems perspective, large language models can be conceptualized as information systems that transform clinical prompts into counseling information, making their reliability critical to evaluate.

Large language models are increasingly used for patient education, and earlier studies suggesting generally accurate sexual health information.^7–9^ Although, LLMs show promise in medicine, their application to counseling women with female sexual dysfunction following cystectomy remains understudied. As interest in FSD within urologic oncology grows, LLMs may serve as tools for patient education. However, LLM-generated counseling may be unreliable or inaccurate for patients experience post-operative sexual dysfunction. LLM chatbots are susceptible to generating incorrect information and often lack nuanced, empathetic counseling for sensitive contexts.^10^ Conversely, recent studies demonstrate that chatbots can deliver sexual health information with high correctness.^11^ Ocakoglu and Coskun found that ChatGPT produced more complex yet more complete outputs than Bing and Google’s BARD.^12^ These findings underscore the need to evaluate whether LLMs can supplement counseling for women with post-cystectomy FSD or risk amplifying post-operative disparities.

Rossettini et al. evaluated ChatGPT, Gemini, Copilot, Claude, and Perplexity against clinical practice guidelines for lumbosacral radicular pain. We selected ChatGPT, Gemini, and Perplexity because they are commonly used and frequently evaluated in prior studies.^13–15^ We evaluated LLM output quality using a custom checklist derived from AUA guidelines for MIBC focused on post-cystectomy sexual health counseling domains.^16^ This approach parallels prior work translating guidelines into structured evaluation benchmarks for assessing LLM-generated responses. ^17^ Current literature in the scope of urology demonstrates that AI models when applied to diagnosis, treatment planning, and outcome prediction, are often superior to conventional statistical methods in terms of accuracy and utility.^18–20^ Furthermore, previous studies have utilized expert panel evaluation of AI chatbot responses to real world sexual health queries.^11^ Several studies have evaluated the reliability, quality, and readability of LLM generated health information. Ozduran et al. examined responses from ChatGPT, Gemini, and Perplexity to the most frequently searched pain related queries.^14^ Similarly, Hanci et al. evaluated five LLMs (ChatGPT, Bard, Gemini, Copilot, Perplexity) on palliative care questions, however they found that readability consistently exceeded recommended health literacy levels and quality scores remained low across models.^15^

The quality and accuracy of the generated content is often the point of focus, yet they are irrelevant in practice if patients cannot comprehend the material. Many studies show that AI-generated materials are too complex for Americans, who on average, read at a sixth-grade level.^21,22^ Despite this, little is known about the methodological underpinnings of readability research. Common formulas used to assess readability include Flesch, Flesch-Kincaid, and SMOG, They provide decent approximations of textual difficulty, and have been shown to be quantitatively correlated as they all rely upon sentence length, word length, and syllable structure.^23,24^ Treating these metrics as independent outcomes (ubiquitous in the literature) inherently increases the probability of type I error, thereby obfuscating model-level differences, and undermining interpretability.^25,26^

To address these limitations, we offer a psychometrically grounded analytical pipeline using principal component analysis and hierarchical clustering; methods previously recommended to reduce multicollinearity among text-based indicators and to identify latent constructs within linguistic datasets.^27,28^ PCA was used to determine whether readability metrics reflect a single dimension or multiple distinct linguistic constructs, ensuring that subsequent inferential models include valid and non-redundant predictors. With this framework, we evaluated LLM-generated sexual health counseling responses following radical cystectomy by assessing guideline adherence and quantifying linguistic complexity across outputs. We hypothesized that both guideline adherence and readability would vary by LLM type and prompt format.

## II. Methods

Due to this study analyzing only AI-generated outputs, it was deemed exempt from review by the Institutional Review Board. We evaluated three LLMs (ChatGPT, Gemini, Perplexity) using two prompt formats (long-form clinical vs short-form lay language).

Responses to six patient questions were generated in separate incognito sessions to minimize cross-prompt carryover.^29^ Two reviewers scored outputs using a 4-item guideline-adherence checklist.

Question inclusion was determined *a priori* by the authors, and based on recommendations from the American Urological Association and the American Society of Clinical Oncology that reflected issues that are typically addressed during pre-operative counseling^16,30^. We chose these 3 LLMs (ChatGPT, Perplexity, and Gemini) due to their accessibility and common usage in associated literature.^13–15^

The dataset consisted of 36 entries [6 Questions x 3 LLMs x 2 Reviewers]. To test the effect of question phrasing, five questions that were used as prompts for the LLMs used technical clinical language in long-form prompts, and one used a lay short-form prompt (**See Supplemental Materials S1**).

Checklist items ranged from [0-4] were transformed to a proportional measure [0-1]. The rubric included: (1) Counseling patients on potential complications and quality of life implications of treatment (sexual function, fertility, bowel dysfunction), (2) Discussing sexual function preserving options for surgery (nerve sparing procedures), (3) Discussing supportive care and mental health counseling for the patient, and (4) Discussing lifestyle modifications to support long-term health and quality of life (smoking cessation, physical activity, balanced diet). Two reviewers evaluated each LLM response, scoring inclusion of each guideline item as “Yes” or “No.” In this context, guideline adherence was utilized as a system-level performance metric reflecting the reliability of LLM-generated counseling outputs.

We purposely presented adherence as coverage of guideline-endorsed counseling domains rather than a comprehensive assessment of sexual medicine expertise. This approach reflects real-world preoperative counseling expectations rather than subspecialty sexual therapy standards.

The primary outcome measure, guideline adherence, was operationalized as the proportion of the guidelines that were satisfied by each LLM response. We employed a linear mixed effects model to predict guideline adherence as a function of LLM type, prompt type, and the reviewer, while accounting for non-independence of responses nested in individual questions. The models all took a similar form:

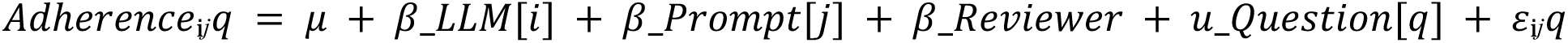

LLM type, prompt type, and reviewer were modeled as fixed effects. We chose to model QuestionID as a random effect to account for clustering and variance in difficulty differences across the questions. Statistical analyses were conducted using R, and model fitting was done using restricted maximum likelihood with the lme4 library using the lmer function and the LmerTest library to calculate p-values using Satterthwaite’s method.^31–33^

If a significant main effect of LLM was found, post-hoc pairwise comparisons of estimated marginal power means (EMMs) using the emmeans package (95% confidence intervals).^34^ Tukey-Kramer correction was applied for pairwise comparisons between LLMs and between prompt formats.^35,36^ Model assumptions were evaluated for normality and homoscedasticity of residuals by inspecting the residual plots and using the performance::check model command. Interrater reliability was assessed with Cohen’s k with k values and assessed with 95% confidence intervals calculated for each LLM.

In an independent analysis, we attempted to model the Reviewer term as a random effect, however, given the near zero variance, we argue it is more sensible as a fixed effect, rather than excluding the term from the model wholly. This is given the randomness of the QuestionID term and given the fixed predictor nature of the Reviewer term. This was confirmed as there was indeed a singularity warning thrown. As suggested by Huo and colleagues, we chose not to model the interaction term [LLM x prompt combination] due to the small number of observations per interaction.^37^

### Readability and Dimensionality Analysis

We quantified the readability of each AI-generated response using five established indices: (1) Flesch Reading Ease, Flesch-Kincaid Grade, ARI, SMOG, and Coleman-Liau index. Scores were computed directly from the full text of each response using the standard formulas for sentence segmentation, word counts, and syllable estimation. To predict textual complexity across models and prompt types, we again used a linear mixed effects framework (equation two). Here the μ represents the grand mean readability score across all responses, βLLM[i] and βPrompt[i] are fixed effects of the language model and prompt type, respectively. μquestion[j] is the random intercept showing the variation due to the clinical question, and εij is the residual error term.

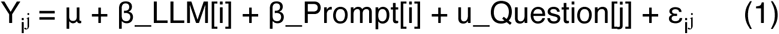

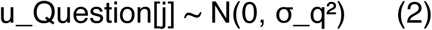

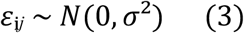

The model estimates the predicted readability score for each response as a function of LLM and prompt type, while also accounting for the shared variance within individual clinical questions. Principal component analysis (PCA) was then applied on z-scored values of all five indices. Missing data were imputed using column means. Component extraction was based on the covariance matrix, and the first two principal components were used to generate a biplot. Hierarchical agglomerative clustering was performed using Ward’s method on a distance matrix defined as 1-r, where r is the Pearson correlation between readability indices. The ordering was applied to the correlation heatmap to show shared structure across the metrics.

All analyses were conducted using R (version 4.5.2) and Python (3.14.0) using the stats, ggplot2, lme4, emmeans, and scikit-learn libraries. The tables corresponding to the aforementioned statistical analyses are in the **Supplementary Tables S2.**

## III. Results

In the following section, we present Its of how guideline adherence and readability metrics vary as a function of LLM-type and prompt type. A total of 36 AI-generated responses were evaluated across three LLMs: ChatGPT, Gemini, and Perplexity. Responses were independently reviewed and scored for adherence to the clinical guidelines regarding post-cystectomy counseling.

We first examined model-level differences in estimated adherence across the three LLMs. A linear mixed effects model showed robust variability in adherence scores as a function of LLM **(Figure 2a).** Tukey-adjusted marginal means show that ChatGPT was the most adherent to the guidelines (emmean =.769, 95% CI [0.645, 0.894]). Gemini was the second most adherent (emmean = 0.499, 95% CI [0.374, 0.623]), with Perplexity in a close third (emmean = 0.457, 95% CI [0.333, 0.581]) (**Table 1**). Pairwise comparisons reinforce the previous finding, showing that ChatGPT outperformed both Gemini (estimate = 0.27, SE = 0.058, *p = 0.0002)* and Perplexity (estimate = 0.31, SE = 0.058, *p* <.0001) (**Table 2**). Forest plots summarizing all fixed-effect coefficients and 95% CIs are presented in **Figure 2**.

Next, we examined the effect of prompt type on adherence, assessing whether framing the question in long-form or short-form influenced adherence scores. We found that when the prompt was phrased in short-form, the LLMs were more likely to elicit higher adherence (emmean = 0.667, 95% CI [0.458, 0.876]), than when more long-form language was used (emmean = 0.483, 95% CI [0.390, 0.577]) **(Figure 2b**). Reviewer identity did not significantly influence adherence score, and the random intercept for question accounted for substantial between-item variability.

We then examined the linguistic complexity of the AI-generated responses. We found a considerable difference between the models. Perplexity generated highly complex, graduate-level prose (μFK Grade ∼16), while Gemini (μFK ∼ 11) consistently produced the lowest grade level content. Interestingly, prompt type did not reduce reading difficulty to a meaningful extent. The shorter prompt produced slightly simplified but still high reading levels.

To evaluate the dimensional structure of the readability space and ensure our modeling strategy was not dependent on a single metric, we conducted a principal component analysis (PCA) of all five standard readability indices. PCA demonstrated that a single dominant component (PC1) accounted for 76.7% of total variance, with Flesch–Kincaid Grade Level, ARI, and the Coleman–Liau Index all loading strongly on this axis (**Figure 1a**). Hierarchical clustering of the correlation matrix showed an identical structure (**Figure 1b**). This pattern makes it clear that the five-readability metrics are all probing the same underlying construct, which supports using Flesch–Kincaid *Grade Level* as the primary outcome in our model. The PCA shows that our findings would not materially change if a different readability index were used, thus strengthening the rigor, stability, and transparency of our modeling approach.

**Figure 1.**
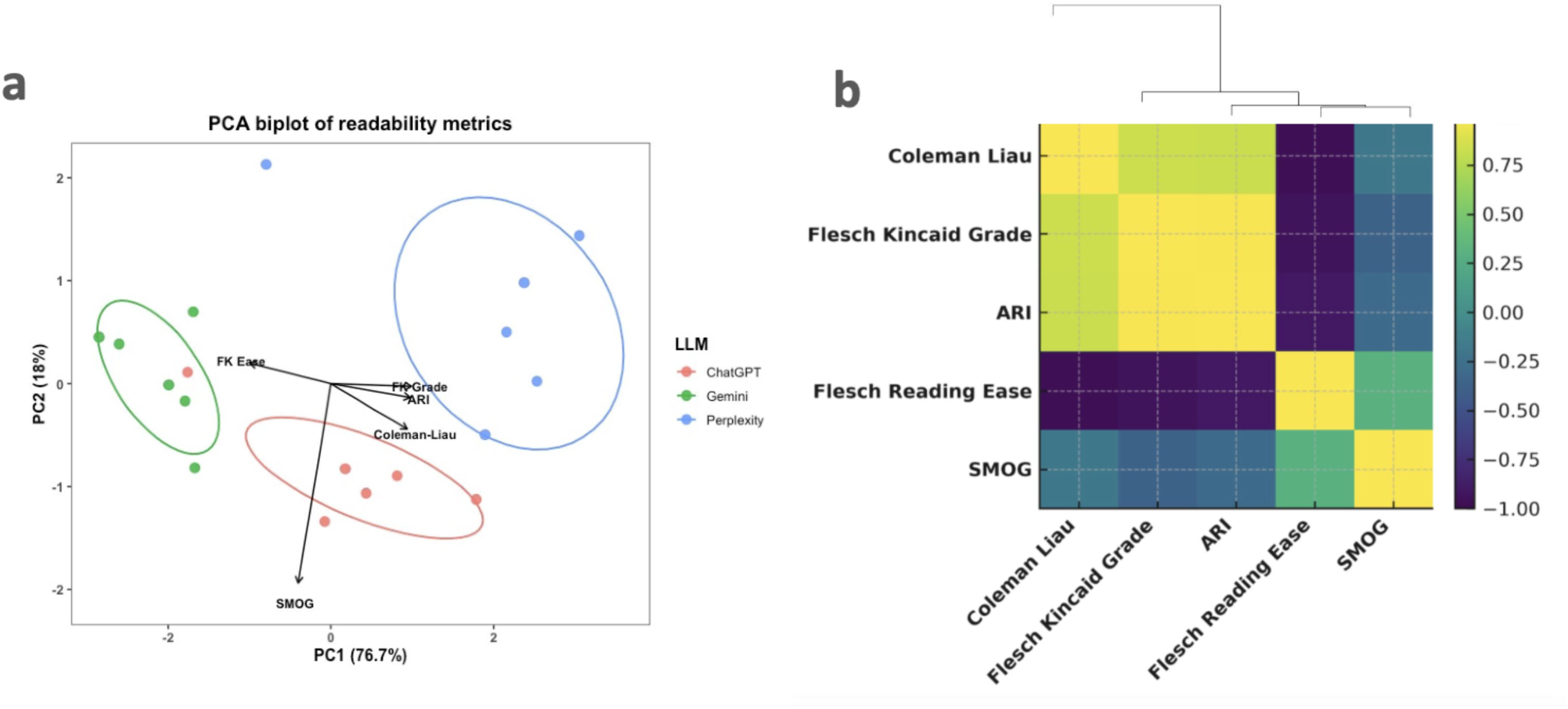
Linguistic complexity across large language model outputs. (a) Principal component analysis (PCA) biplot of five readability metrics [Flesch Kincaid Grade Level, Flesch Reading Ease, SMOG, Coleman-Liau Index, and Automated Readability Index] applied to responses generated by ChatGPT, Gemini, and Perplexity. Each point represents an individual model output. Vectors indicated the contribution and directionality of each readability metric to the principal components. (b) Hierarchical clustering heatmap representing pairwise correlations between readability metrics across all outputs. Strong positive correlations (yellow) and negative correlations (purple).

**Figure 2.**
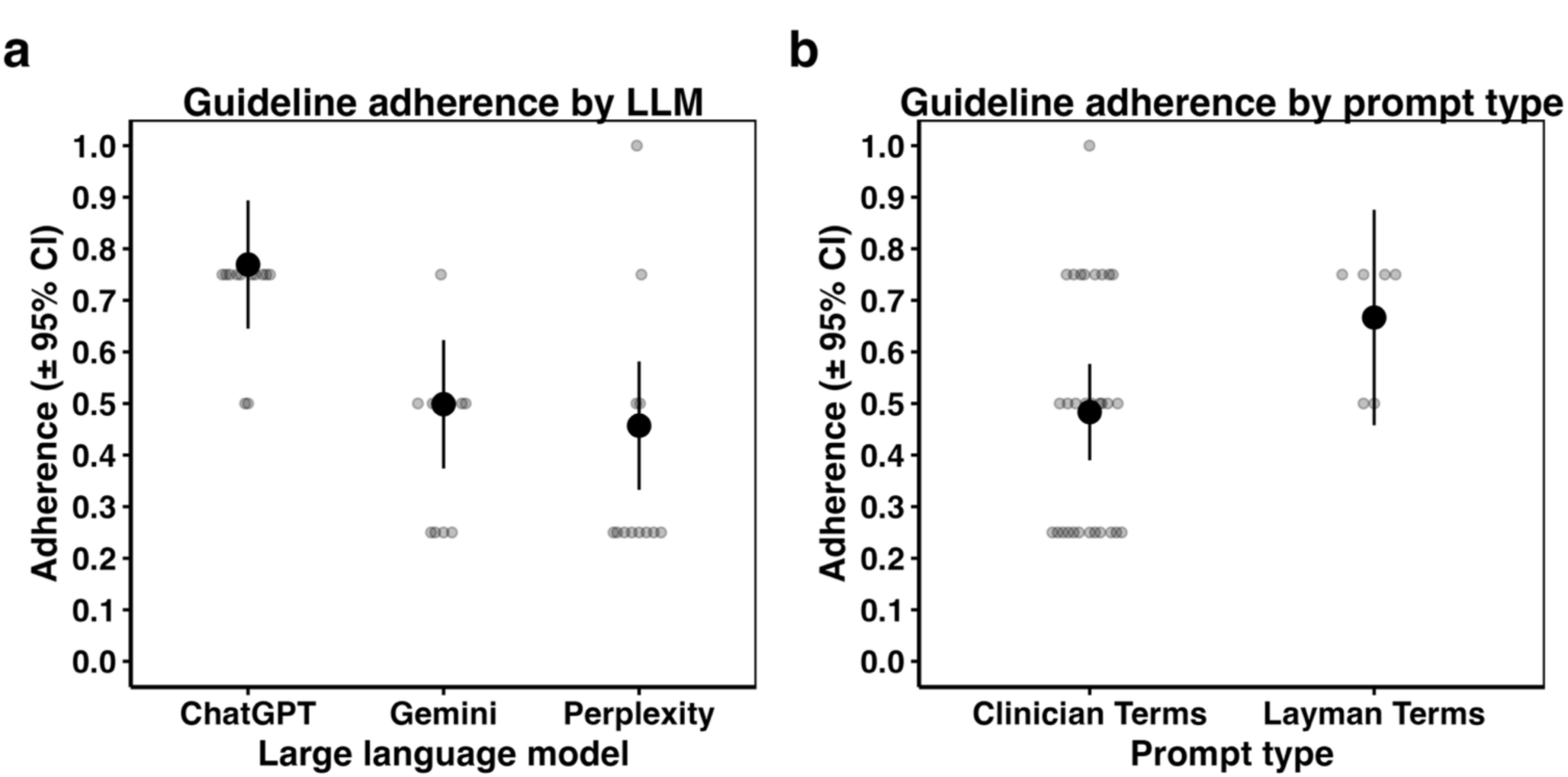
Guideline Adherence scores across large language models and prompt types. (a) Mean adherence to American Urological Association (AUA) guideline components by large language model (ChatGPT, Gemini, Perplexity). Each black point represents the model’s mean adherence score across all questions, with vertical lines indicating 95% confidence intervals. Individual question-level adherence scores are shown as grey points. (b) Mean guideline adherence stratified by prompt type, short-form (layman-worded) vs long-form (clinician-worded) prompts. Black points denote mean adherence with 95% confidence intervals, and grey points represent individual adherence values

**Figure 3.**
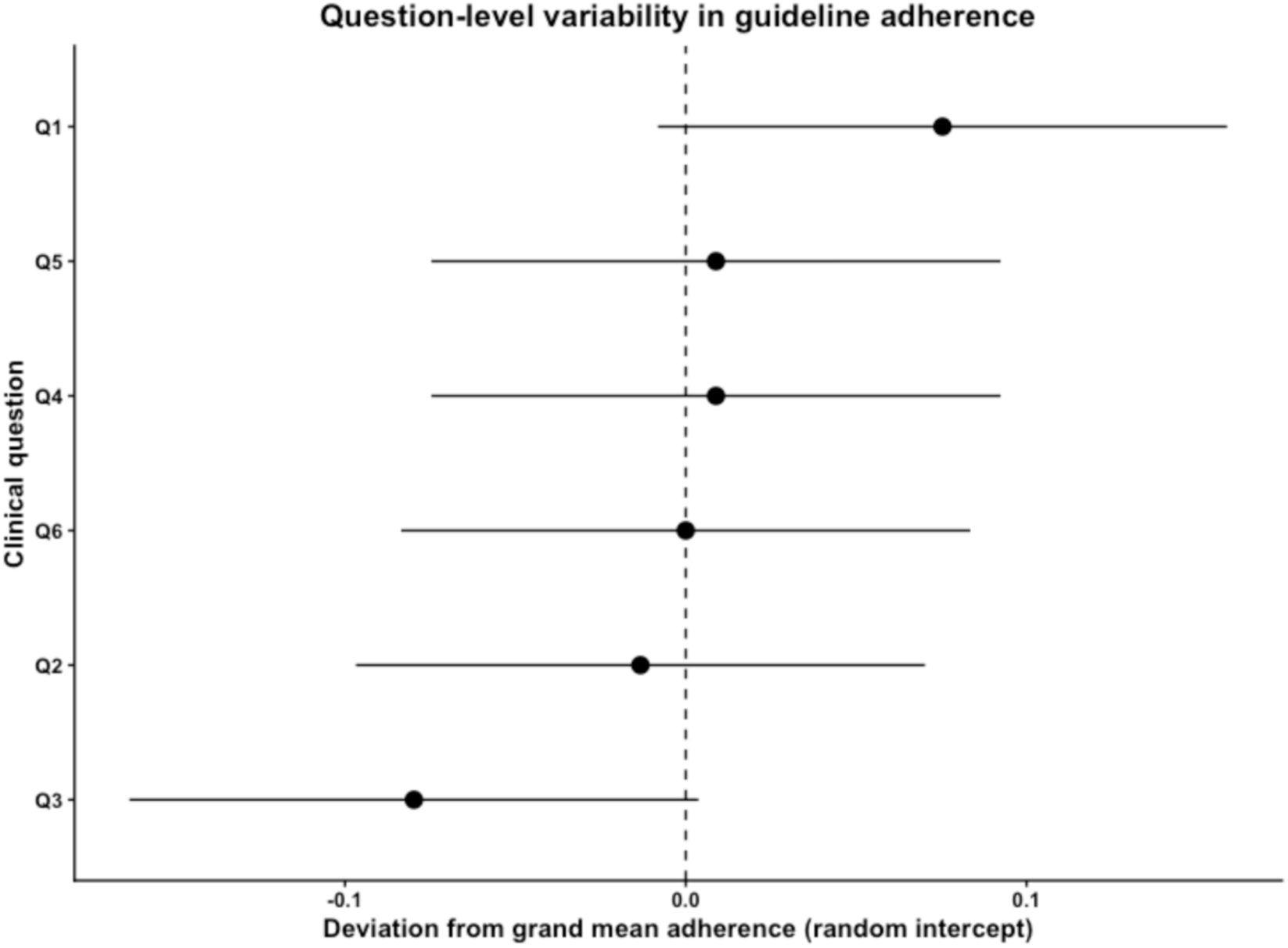
Question level variability in guideline adherence. Random intercepts for each clinical question from the linear mixed model predicting adherence. Points represent estimated deviations from the grand mean adherence, and horizontal lines denote 95% confidence intervals. Values to the right of zero indicate questions that elicited higher-than-average adherence across models. Values to the left indicate systematically under-covered topics. For full length questions are available upon request.

**Figure 4.**
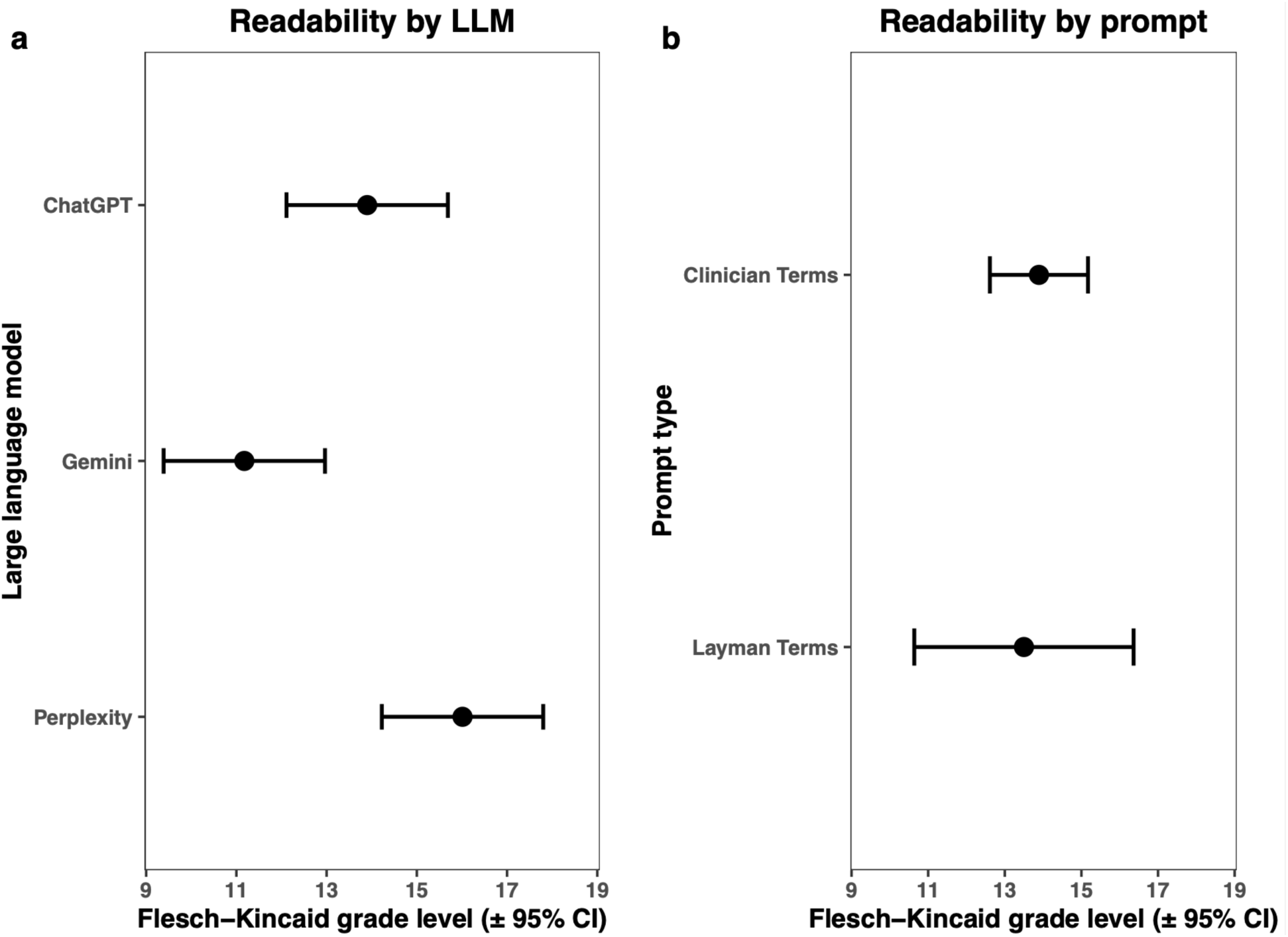
Readability of LLM-generated responses by model and prompt type. Estimated marginal means (±95% CI) of Flesch–Kincaid Grade Level are shown for each large language model (ChatGPT, Gemini, and Perplexity) (panel A) and for clinician-oriented vs lay-oriented prompts (panel B). Higher values indicate more complex reading levels. ChatGPT consistently produced responses at lower grade levels than Gemini or Perplexity, and lay-style prompts yielded substantially lower reading difficulty compared with clinician-style prompts.

## IV. Discussion

In this study, we found that when evaluating LLMs as medical information systems, ChatGPT demonstrated the highest compliance with AUA guidelines in addressing patient questions regarding female sexual dysfunction following radical cystectomy. Additionally, ChatGPT performed significantly better when compared to Gemini and Perplexity, whereas no significant difference was observed between the latter two models. Finally, there was a significant effect of prompt type, such that shorter, lay-style prompts generated more guideline-aligned content than long, clinician-worded prompts, suggesting that conversational framing can meaningfully shape AI outputs in this domain.

Our findings align with prior work that suggests that LLMs deliver uneven quality content in sexual health and oncologic contexts, with performance differences across task complexity and model architecture.^38–42^ In general, studies within the realm of urology show that ChatGPT produces accurate content, but is often inconsistent in the completeness and specificity. This effect is particularly magnified in more nuanced clinical topics, specifically LUTS/BPE counseling, prostate cancer management, and urolithiasis education.^43–45^ Prior evaluations of AI generated sexual health information similarly report inconsistent correctness and actionability, particularly for psychosocially complex queries.^11^ These findings reinforce that model selection matters and even high performing LLMs may produce incomplete counseling content in sensitive contexts.

The bulk of relevant literature focuses purely on clinical accuracy, and to build on this, we evaluated the LLMs’ ability to adhere to *patient-centered* counseling elements derived from AUA/ASCO guidelines for MIBC. Short, lay-language prompts generated higher guideline adherence than clinician-worded prompts. Consistent with prior studies, clinical-style prompts may favor precise medical reasoning.^46^ Informal prompts tend to generate broader patient-centered counseling elements.^47^ Our rubric emphasized mental health support, lifestyle counseling, and discussion of sexual function/post-operative quality of life. Together, these findings suggest that prompt design is a critical determinant of guideline adherence.

Limited readability of LLM-generated content is a barrier to clinical translation and risks exacerbating health disparities. Many studies demonstrate that AI-generated content exceeds recommended reading levels, particularly among outputs generated by Perplexity.^15,21^ This limitation is especially consequential for underserved populations.^48,49^ Providing these patients with unexamined LLM-generated responses could worsen disparities which has been echoed by many scholars.^50–52^ An important consideration indeed, as gender inequities exist in bladder-cancer evaluation, surgical counseling, and survivorship care for women.^5,53^

Readability metrics provide an indirect index of cognitive burden imposed on patients attempting to comprehend information at a vulnerable time. Psychosocial distress post-cystectomy represents a determinant of survivorship quality of life. Body image distress is especially relevant in women undergoing urinary diversion, with evidence demonstrating persistent disparities despite partial functional recovery over time.^54,55^ Studies often demonstrate that women experience lower emotional, mental health, and sexual functioning, compared with men, with these disparities being consistent over time.^4^ Psychosocial responses to urologic diagnoses and procedures frequently include mental distress and burden during postoperative adaptation.^56^ Sexual dysfunction and mental health are related, women describe disappointment with early sexual experiences, avoidance of intimacy, and heightened sexual anxiety due to psychological and physical limitations.^57^ Despite this, sexual health counseling is undervalued, with female patients less likely than males to receive counseling on sexual function, psychological well-being, and survivorship needs.^5,6^ Collectively, this literature underscores that postoperative sexual health in women is inseparable from mental health, psychosocial support, and accessibility of counseling that directly contextualize the evaluation of AI-mediated patient education tools.

Ethical concerns surrounding accuracy, bias, privacy, and inability to hold LLMs accountable for their actions remain paramount.^58–60^ LLMs may generate incomplete or emotionally insensitive responses and lack the capacity to provide individualized, trauma-informed counseling for women navigating sexual dysfunction following cancer treatment.^37,61^ While regulatory and professional guidance has begun to address transparency and disclosure in AI-assisted clinical tools, responsibility for appropriate use ultimately rests with clinicians. Physician should remain aware of ethical standards and professional guidelines governing AI in medicine, as reliance on unvetted AI counseling content creates liability. These conditions emphasize the need for cautious, clinician-supervised deployment of LLMs in sensitive counseling contexts.

Our study is not without limitation. First, we assessed only adherence to guideline components, not the factual accuracy or tone of LLM responses. A greater adherence score does not necessarily imply high communication quality. We had a much smaller sample size of LLM outputs (N=6) that were prompted with short-form lay-language, and a small sample size in general, thus limiting the generalizability of the associated findings. Finally, our checklist was derived from AUA guidelines on muscle-invasive bladder cancer, and not specifically on guidelines related to female sexual dysfunction. Although the guidelines are validated by urologists, this checklist may not encompass all nuanced aspects of postoperative counseling. Future work should also focus on increasing the number of model comparators, increasing the number of both types of prompts, and investigating male sexual dysfunction.

## V. Conclusion

In sum, we demonstrate that the type of LLM matters when counseling for female sexual dysfunction following radical cystectomy, with ChatGPT demonstrating the strongest adherence. Additionally, we saw a considerable linguistic complexity difference between each model. Future research should evaluate whether this performance generalizes across other urologic conditions and patient populations. Our study demonstrates that LLMs can provide equitable and accurate support for patient understanding of sexual health after major urological surgery, while also laying the groundwork for future research by illustrating how more sophisticated statistical approaches can enhance data representation and support stronger conclusions.

## Supporting information

Supplementary Questions

## Data Availability

All data produced in the present study are available upon reasonable request to the authors

