## Supplementary Questions for "Evaluation of Large Language Models for Post-Cystectomy Sexual Health Counseling in Women"

**Supplementary Materials S1**

Q1: As a woman about to undergo Radical cystectomy for Muscle-Invasive Bladder Cancer, Will I be able to have sex after?  Please ensure the information is medically accurate and based on the current best practices/guidelines for urology/urogynecology. Please use clinical language while ensuring the language is understandable by a layperson. Include images and diagrams if applicable.

Q2: As a woman about to undergo Radical cystectomy for Muscle-Invasive Bladder Cancer, What kind of changes will I experience in regards to orgasms after cystectomy for Bladder Cancer?  Please e snsure the information is medically accurate and based on the current best practices/ guidelines for urology/urogynecology. Please use clinical language while ensuring the language is understandable by a layperson. Include images and diagrams if applicable.

Q3: As a woman about to undergo Radical cystectomy for Muscle-Invasive Bladder Cancer, What will my partner feel during sexual intercourse after my radical cystectomy for Bladder Cancer?  Please ensure the information is medically accurate and based on the current best practices/ guidelines for urology/urogynecology. Please use clinical language while ensuring the language is understandable by a layperson. Include images and diagrams if applicable.

Q4: As a woman about to undergo Radical cystectomy for Muscle-Invasive Bladder Cancer What things can I do to maintain sexual function after Radical cystectomy for Bladder Cancer?  Please ensure the information is medically accurate and based on the current best practices/ guidelines for urology/urogynecology. Please use clinical language while ensuring the language is understandable by a layperson. Include images and diagrams if applicable.

Q5: As a woman about to undergo Radical cystectomy for Muscle-Invasive Bladder Cancer, Will I experience pain with sex after radical cystectomy for Bladder Cancer?  Please ensure the information is medically accurate and based on the current best practices/ guidelines for urology/urogynecology. Please use clinical language while ensuring the language is understandable by a layperson. Include images and diagrams if applicable.

Q6: I am a woman with bladder cancer and I am getting my bladder removed. How will this affect my sex life?
