## Supplementary Tables for "Evaluation of Large Language Models for Post-Cystectomy Sexual Health Counseling in Women"

**Table 1:** Estimated marginal means of adherence by large language model (LLM).

| LLM | Mean | Lower 95% CI | Upper 95% CI |
|---|---|---|---|
| ChatGPT | 0.769 | 0.645 | 0.894 |
| Gemini | 0.499 | 0.374 | 0.623 |
| Perplexity | 0.457 | 0.333 | 0.581 |

**Table 2:** Tukey-adjusted pairwise comparisons of adherence between LLMs.

| Contrast | Estimate | SE | df | t | p-value |
|---|---|---|---|---|---|
| ChatGPT – Gemini | 0.271 | 0.0584 | 30 | 4.635 | 0.0002 |
| ChatGPT – Perplexity | 0.313 | 0.0584 | 30 | 5.348 | < 0.0001 |
| Gemini – Perplexity | 0.042 | 0.0584 | 30 | 0.713 | 0.758 |

**Table 3:** Estimated marginal means of adherence by prompt type.

| Prompt type | Mean | Lower 95% CI | Upper 95% CI |
|---|---|---|---|
| Long article | 0.483 | 0.390 | 0.577 |
| Short question | 0.667 | 0.458 | 0.876 |

**Table 4:** Estimated marginal means of readability (Flesch–Kincaid grade level) by LLM.

| LLM | Mean grade level | Lower 95% CI | Upper 95% CI |
|---|---|---|---|
| ChatGPT | 14.03 | 12.35 | 15.71 |
| Gemini | 11.31 | 10.23 | 12.38 |
| Perplexity | 16.14 | 13.71 | 18.58 |
